# Factors Associated with Seeking and Receiving Home Antiviral Therapy During the COVID-19 Pandemic

**DOI:** 10.1101/2025.05.25.25328310

**Authors:** Stuart F. Quan, Matthew D. Weaver, Mark É. Czeisler, Lauren A. Booker, Mark E. Howard, Melinda L. Jackson, Christine F. McDonald, Anna Ridgers, Rebecca Robbins, Prerna Varma, Shantha M.W. Rajaratnam, Charles A. Czeisler

## Abstract

**Background:** Home antiviral treatment (HAVT: nirmatrelvir/ritonavir [Paxlovid] and molnupiravir [Lagevrio]) for COVID-19 was approved in December 2021 by the U.S. Food and Drug Administration, but ensuing utilization during the COVID-19 pandemic was low. Factors associated with seeking HAVT treatment have not been fully explored.

**Study Design:** Cross-sectional survey of 9,944 U.S. adults to identify factors associated with seeking and receiving HAVT in persons with confirmed or suspected COVID-19.

**Results:** COVID-19 infection was confirmed or suspected in 4,355 (43.8%) participants. HAVT was sought by 1,331 (26.3%) and medication was received by 928 (20.7%). In comparison to those who were COVID-19 test negative and had no loss of taste or smell (aOR: 1.856, 95% C.I.: 1.262-2.728), the factor most associated with seeking HAVT was a loss of taste or smell irrespective of a positive (aOR: 3.823, 95% C.I.: 2.626-5.567) or negative COVID-19 test (aOR: 3.306; 95% C.I.: 2.086-5.241). Loss of taste or smell was similarly associated with receiving HAVT. Male sex, younger age, Black race, Hispanic ethnicity, conservative or liberal political preference, and medical discrimination were some of the other factors associated with a greater likelihood of seeking HAVT. Barriers to obtaining treatment included feeling uncomfortable obtaining HAVT while sick and lack of transportation; 14.7% of those seeking treatment reported at least one barrier.

**Conclusion:** In a general population, HAVT was an underutilized resource during the COVID-19 pandemic. Loss of taste or smell was the most important factor among several others associated with seeking HAVT irrespective of COVID-19 test status.

## Introduction

On December 22, 2021, Emergency Use Authorization (EUA) was granted by the United States Food and Drug Administration (FDA) for nirmatrelvir/ritonavir (Paxlovid) as the first oral home antiviral treatment (HAVT) for COVID-19. This was followed the next day by an EUA for a second HAVT, molnupiravir (Lagevrio). Patient groups initially selected for priority use were adults and children (older than 12 years and weighing at least 40 kg) with mild to moderate COVID-19 who were at high risk for progression to severe disease (e.g., compromised immunity, lack of vaccination, age >75 years or age <65 years with chronic medical conditions)[1]. Final FDA approval for nirmatrelvir/ritonavir in adults was granted on March 13, 2024 although the EUA for molnupiravir and for nirmatrelvir/ritonavir in children remains in effect. Use of HAVT in adults now includes those with mild to moderate COVID-19 who are at high risk for progression to severe disease [2].

As availability of HAVT increased, programs such as “Test to Treat” to promote public awareness of HAVT as well as efforts to educate health care providers were initiated [3]. These included a change in regulation to allow pharmacists to prescribe nirmatrelvir/ritonavir [4]. Efforts were made to ensure equitable distribution of medication to populations with reduced access to healthcare [5]. Nevertheless, despite evidence demonstrating that nirmatrelvir/ritonavir and to a lesser extent molnupiravir are effective in treating COVID-19 [6,7], data suggest public awareness of nirmatrelvir/ritonavir is low [8] and there is underutilization, with prescriptions dispensed in less than 25% of eligible patients [9–13]. Furthermore, although these HAVTs for COVID-19 were initially available at no cost to patients, studies indicate that during the first year of availability, there were substantial disparities in dispensing rates between low and high social vulnerability areas [14]. Several studies documented that individuals who were African American or Hispanic were prescribed HAVT at lower rates than those who were Non-Hispanic White [11,15,16]. In most studies, these data were obtained from electronic health records (EHRs) used by networks of pharmacies and healthcare systems or medical claims databases [9–12,14,16,17]. The only previous community based survey on characteristics associated with HAVT-seeking behaviors was performed in Singapore, where indications included age 18 years or older, presentation within 5 days of illness onset and at risk of developing severe disease. The study found that the intention to seek a nirmatrelvir/ritonavir prescription was associated with male sex, pre-existing diabetes and perceived severity of COVID-19 [18]. However, there are no studies in a more diverse population that have examined a broader spectrum of factors associated with seeking and/or receiving a prescription for HAVT.

Mistrust in healthcare has been identified as a factor in COVID-19 vaccine hesitancy and uptake [19,20]. Furthermore, individuals who have experienced discrimination in healthcare settings are also less likely to be vaccinated against COVID-19 [21]. Whether such concerns reduce the likelihood of seeking HAVT for COVID-19 has not been previously assessed.

The purpose of this analysis was to ascertain in a general United States population cohort demographic, socioeconomic, and comorbid medical factors associated with seeking a prescription for a COVID-19 HAVT as well as to identify factors associated with receiving one. Additionally, we assessed whether healthcare mistrust and medical discrimination affected a decision to seek HAVT. To accomplish these goals, we utilized data from 4th and 5th waves of The COVID-19 Outbreak Public Evaluation (COPE) Initiative (http://www.thecopeinitiative.org/), which focused on accumulating data on public attitudes, behaviors and beliefs related to the COVID-19 pandemic from large scale, demographically representative samples.

## Methods

### Study Design and Participants

From March 10, 2022 to October 15, 2022, the COPE Initiative administered five successive waves of surveys focused on accumulating data on the prevalence and sequelae of COVID-19. Questions concerning the use of oral antiviral medications for COVID-19 infection were asked on the 4^th^ wave (August 1-18, 2022) and 5^th^ wave (September 26-October 15, 2022) and form the basis of this analysis; each wave consisted of more than 5000 unique participants who were recruited to approximate population estimates for age, sex, race, and ethnicity based on the 2020 U.S. Census. Surveys were conducted online by Qualtrics, LLC (Provo, Utah, and Seattle, Washington, U.S.), using their network of participant pools with varying recruitment methodologies that include digital advertisements and promotions, word-of-mouth and membership referrals, social networks, television and radio advertisements, and offline mail-based approaches. Informed consent was obtained electronically. The study was approved by the Monash University Human Research Ethics Committee (Study #24036).

### Survey Items

Each survey contained identical items related to COVID-19 infection status and the number of COVID-19 vaccinations participants had obtained. Ascertainment of past COVID-19 infection was obtained using responses from the following questions related to COVID-19 testing or the presence of loss of taste or smell based on a question modified from the Sino-nasal Outcome Test (SNOT-22):

1. “Have you ever tested positive?”
2. “Despite never testing positive, are you confident that you have had COVID-19?”
3. “Despite never testing positive, have you received a clinical diagnosis of COVID-19?”
4. “Have you experienced a problem with decreased sense of smell or taste at any point since January 2020?”

Additionally, participants were asked if they had been exposed to someone who had COVID-19, if they were a member of a group at high risk for COVID-19 or if they were experiencing symptoms concerning for COVID-19 (e.g., fever, tiredness, dry cough, shortness of breath, aches and pains, sore throat).

COVID-19 vaccination status was ascertained by asking “How many COVID-19 vaccine doses have you received?”. Participants were allowed to respond from 0 to 4.

Each survey included the following questions related to potential use of oral antiviral medications for COVID-19:

1. Did you seek to obtain at-home treatment for COVID-19? (for example, Paxlovid or Molnupiravir)?
2. Were you prescribed an at-home treatment for COVID-19? (for example, Paxlovid or Molnupiravir)?
3. Were you able to obtain the treatment?

On the Wave 5 survey, participants were asked whether they received a prescription for nirmatrelvir/ritonavir or molnupiravir.

Participants self-reported demographic, anthropometric, and socioeconomic information including age, race, ethnicity, sex, height and weight, education level, employment status and household income. In addition, they reported information on several current and past medical conditions by answering the question: “Have you ever been diagnosed with any of the following conditions?” An opportunity was provided to endorse high blood pressure, cardiovascular disease (e.g., heart attack, stroke, angina), gastrointestinal disorder (e.g., acid reflux, ulcers, indigestion), cancer, chronic kidney disease, liver disease, sickle cell disease, chronic obstructive pulmonary disease, asthma, and insomnia. Possible responses to each medical condition were “Never”, “Yes I have in the past, but don’t have it now”, “Yes I have, but I do not regularly take medications or receiving treatment”, and “Yes I have, and I am regularly taking medications or receiving treatment”. As defined in several other analyses from the COPE Initiative, the presence of obstructive sleep apnea (OSA) was estimated using a combination of responses to items related to snoring, breathing pauses, and sleepiness [22]. As documented in Supplement 1, information also was obtained regarding health insurance coverage, political beliefs, hours worked per week, night shift work, and whether the participant was a healthcare worker.

On the Wave 4 survey, distrust in healthcare was ascertained using the Health Care System Distrust Scale [23]. Also only on the Wave 4 survey, information regarding health care discrimination was obtained using the Discrimination in Medical Settings Scale [24].

### Statistical Analyses

After preliminary analyses, we limited our examination to only participants who endorsed testing positive for COVID-19, received a clinical diagnosis of COVID-19, were confident they had COVID-19 despite not having a positive test or clinical diagnosis, or who had lost their taste or smell. To assess the combined impact of COVID-19 testing and the presence or absence of the loss of taste or smell, participants were assigned to one of four groups based on COVID-19 test and symptoms: 1) COVID-19 test positive and loss of taste or smell; 2) COVID-19 test positive and no loss of taste or smell; 3) COVID-19 test negative and loss of taste or smell; and 4) COVID-19 test negative and no loss of taste or smell. Seeking HAVT for COVID-19 was defined as an affirmative response to either nirmatrelvir/ritonavir or molnupiravir.

Participants were considered to have OSA if they endorsed currently having the condition whether treated or not or if they had two or more symptoms of OSA (i.e., snoring, breathing pauses, or sleepiness) [22]. Insomnia was present if endorsed by participants irrespective of treatment status [25]. Vaccination status was dichotomized as Boosted (>2 vaccinations) or Not Boosted (≤2 vaccinations) [22]. Comorbid medical conditions were defined as currently having the condition whether treated or untreated. As done in previous COPE analyses, the effect of comorbid medical conditions was evaluated by summing the number of conditions reported by the participant (minimum value 0, maximum value 9) [22,25]. Body mass index (BMI) was calculated using self-reported height and weight as kg/m^2^. Socioeconomic covariates were dichotomized as follows: employment (full or part-time, student vs. retired), healthcare provider (Yes, irrespective of treatment of COVID-19 patients vs. Not a healthcare worker), education (high school or less vs. some college) and annual household income in U.S. Dollars (<$50,000 vs ≥$50,000). Political preference was collapsed into 3 categories: Liberal, Moderate and Conservative. Night shift work was defined as occurring more than 1-2 times per week.

Summary data for continuous variables are reported as their respective means and standard deviations (SD) and for categorical variables as their percentages. Comparisons of co-morbid medical, demographic, and social characteristic variables stratified by whether or not participants sought home HAVT were performed using Student’s unpaired t-test for continuous variables and χ^2^ for categorical variables.

Multivariate modelling using logistic regression was utilized to determine whether seeking HAVT was associated with categories of COVID-19 test and symptoms after controlling for demographic, anthropometric, socioeconomic factors, and comorbidities. Inasmuch as assessments of medical discrimination and distrust in healthcare were only available on the Wave 4 survey, two models were developed. One used data from both waves but excluded evaluation of medical discrimination and mistrust in healthcare. The other included the latter two factors but used only Wave 4 data. Multiple imputation with the use of chained equations was performed separately in each model to address missing data under a missing-at-random assumption. For both analyses, a baseline model was constructed using only COVID-19 test and symptoms data. We then developed fully adjusted models by including demographic, anthropometric, socioeconomic factors, and comorbid medical conditions found to be associated in the baseline models. Additionally, a logistic regression model was developed to assess the association of receiving HAVT with demographic, anthropometric, socioeconomic factors and comorbid medical conditions. Results of the logistic regression models are presented as unadjusted or adjusted odds ratios (aOR) and their 95% confidence intervals (95% CI).

All analyses were conducted using IBM SPSS version 28 (Armonk, NY). A p<0.05 was considered statistically significant.

## Results

As displayed in the Figure, of the 9,944 participants in Waves 4 and 5, 4,355 (43.8%) endorsed testing positive for COVID-19, receiving a clinical diagnosis of COVID-19, were confident they had COVID-19 despite not having a positive test or clinical diagnosis, or had lost their taste or smell. Home antiviral treatment was sought by 1,331 (30.6%) and medication was ultimately received by 928 (69.7% of those who sought HAVT and 21.3% who had or may have had COVID-19). Of those who received COVID-19 HAVT, 69.4% received nirmatrelvir/ritonavir and 30.6% received molnupiravir.

Table 1 shows the prevalence of the 4 groups of COVID-19 test and symptoms as well as various demographic, anthropometric, and comorbid characteristics of the cohort stratified by whether or not participants sought HAVT. Participants who had lost their taste or smell, irrespective of their COVID-19 test results, were more likely to seek HAVT than those who tested positive without loss of taste or smell and those who neither tested positive nor loss their taste or smell. Additionally, male sex, Black race, Hispanic ethnicity and living with a partner were associated with a greater likelihood of seeking HAVT. Higher rates of seeking HAVT were observed in those with comorbid conditions including OSA and insomnia, with exposure to COVID-19, who were members of a COVID-19 high risk group, and who had symptoms suggestive of COVID-19 other than loss of taste or smell. Socioeconomic factors related to seeking HAVT treatment were higher income and level of education, full or part-time employment or student, longer work hours and having health insurance. Both liberal and conservative political preference were associated with a greater likelihood of HAVT. Those who distrusted healthcare or who experienced medical discrimination also had greater rates of seeking HAVT.

**Table 1:**
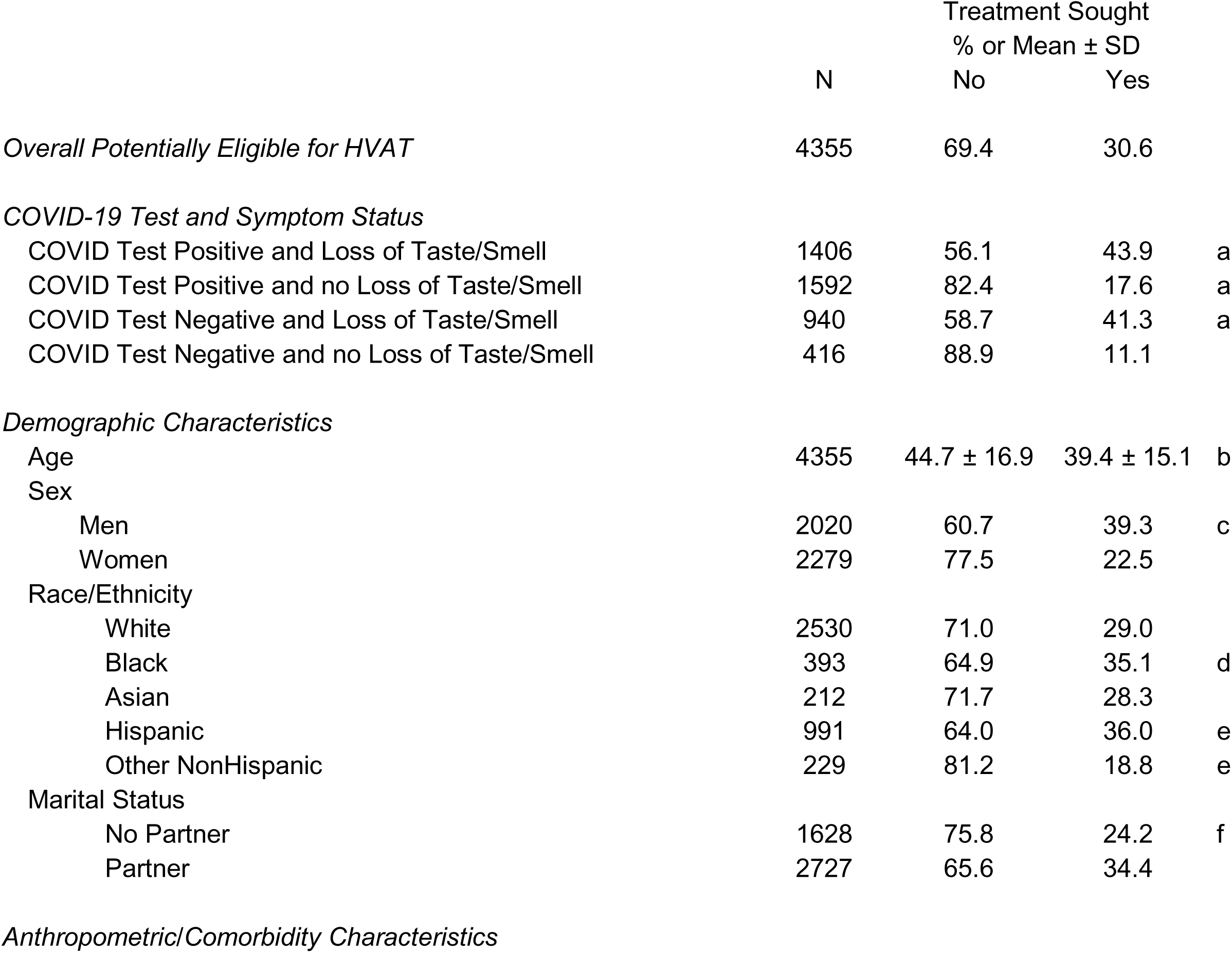
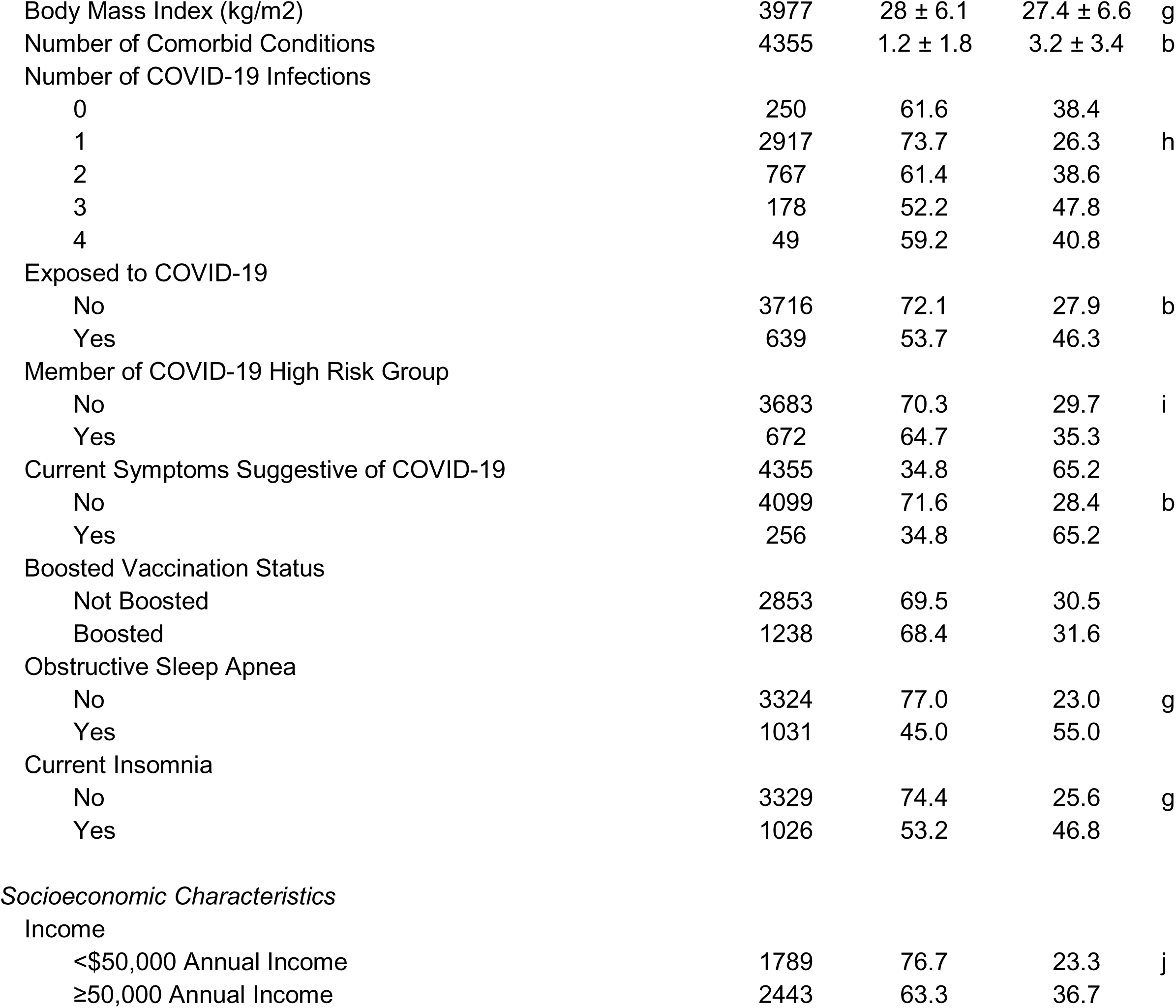
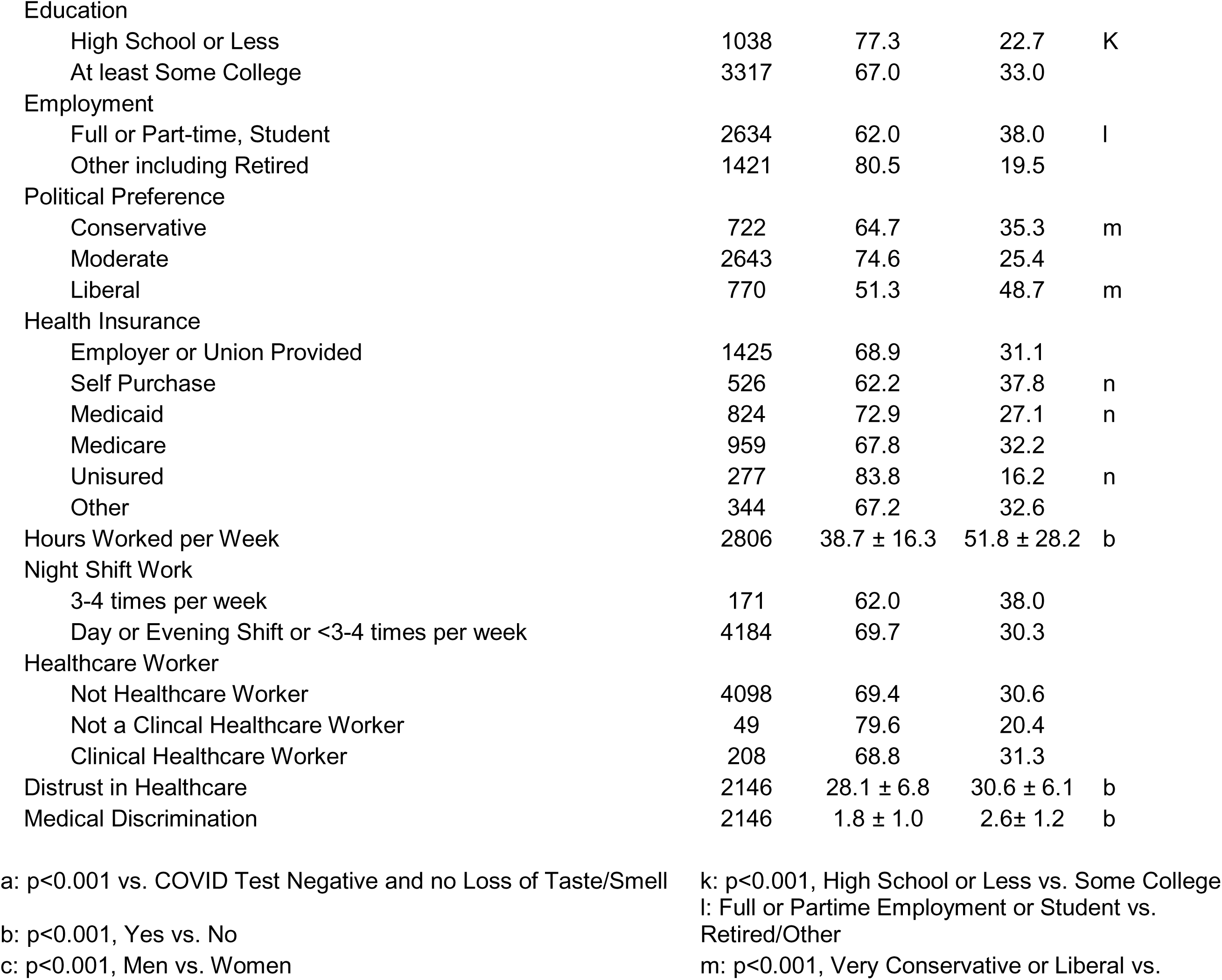
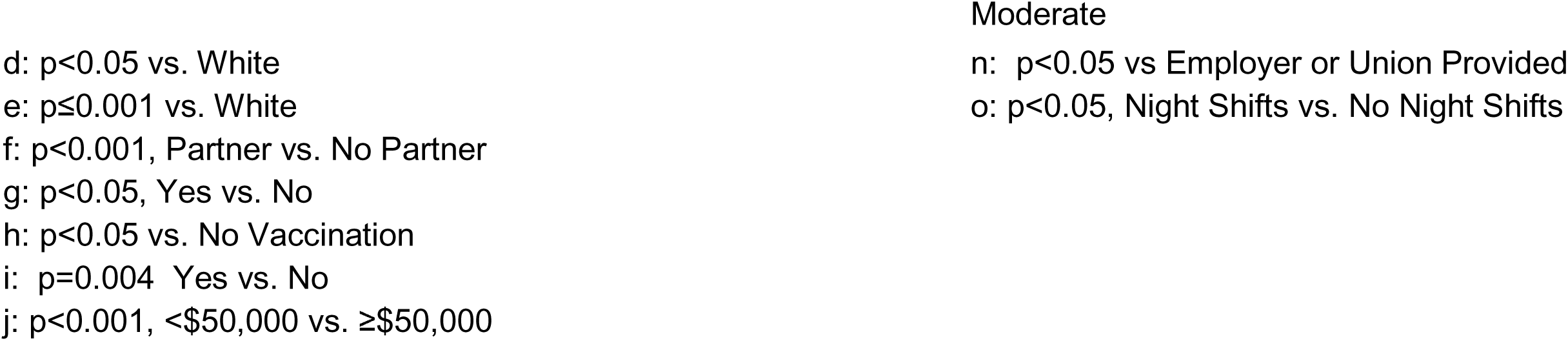
Demographic, Anthropometric and Socioeconomic Characteristics Associated with Seeking Home Antiviral Treatment.

A baseline model showing the unadjusted odds ratios (OR) of COVID-19 test and symptoms and other factors potentially related to seeking HAVT are shown in the Supplemental Table S1. Several demographic, socioeconomic and medical comorbid factors including COVID-19 test and symptoms were linked to seeking HAVT. Displayed in Table 2 are the aOR and 95% CI of factors associated with seeking HAVT for COVID-19 excluding evaluation of distrust in healthcare and medical discrimination. In comparison to those who did not have a positive COVID-19 test and who did not have loss of taste or smell, those who had a positive COVID-19 test and had lost their taste or smell were ∼3.8 times as likely to have sought HAVT (aOR: 3.823, 95% CI: 2.626-5.567). Those who had a negative COVID-19 test but had lost their taste or smell also had a high probability of seeking HAVT (aOR: 3.306, 95% CI: 2.086-5.241). In comparison, although having only a positive COVID-19 test did increase the likelihood of pursuing HAVT (aOR: 1.856, 95% CI: 1.262-2.728), it was less in comparison to the groups that had lost their taste or smell (positive COVID-19 test/loss of taste or smell aOR: 3.823, 95% CI: 2.626-5.567; negative COVID-19 test/loss of taste or smell aOR: 3.307, 95% CI: 2.086-5.241).

**Table 2:**
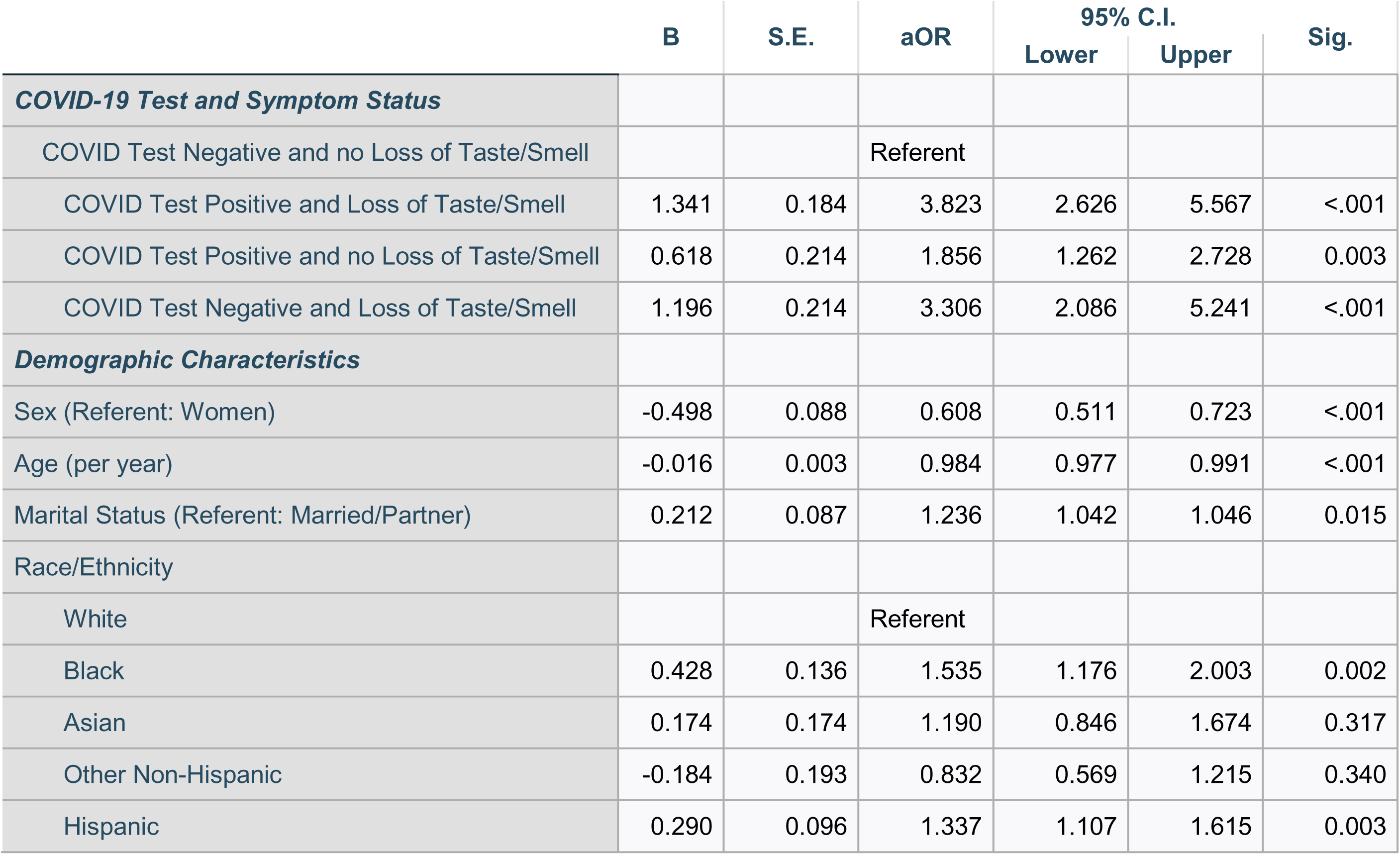
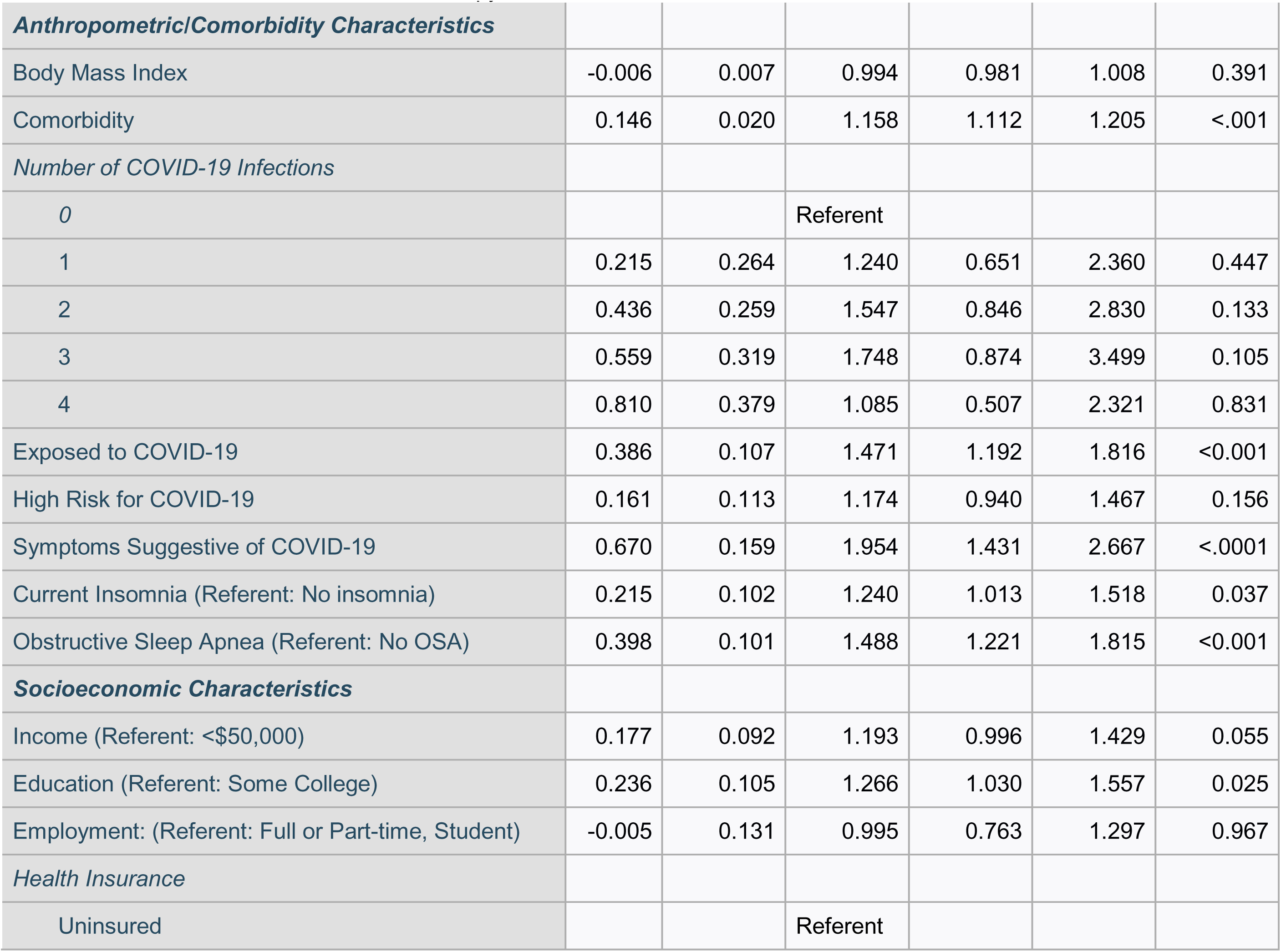
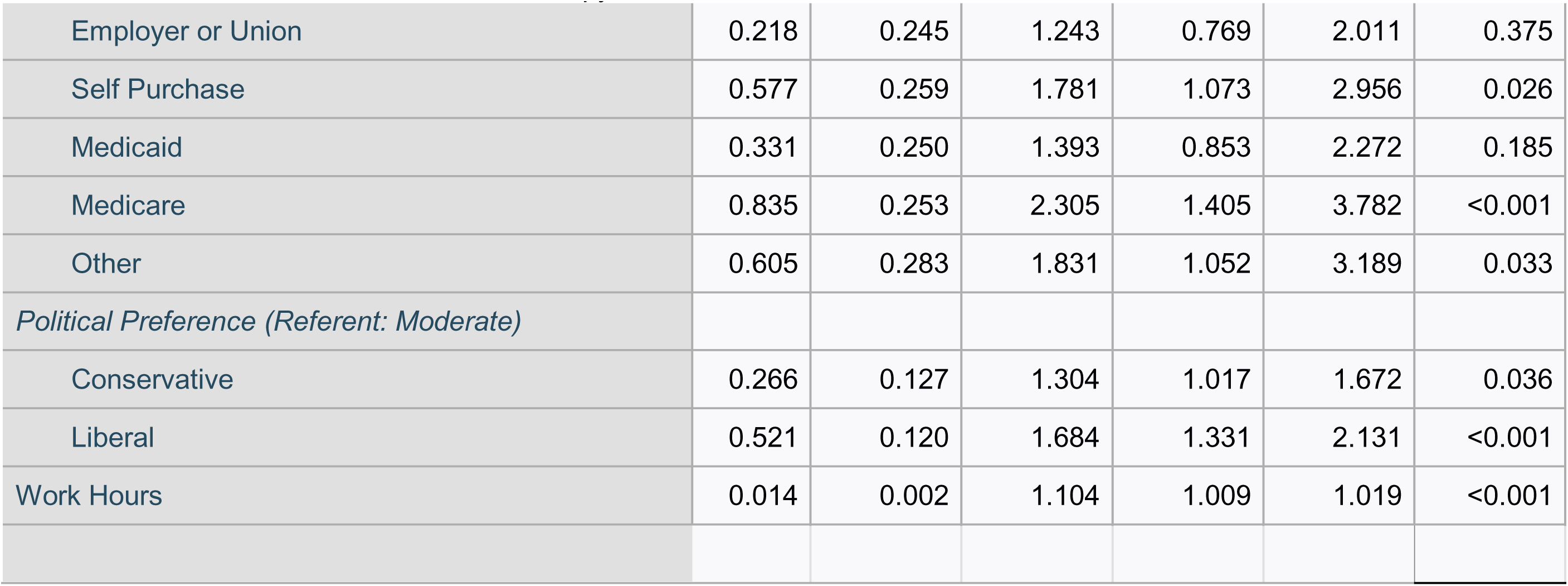
Adjusted Odds Ratios of Factors Associated with Seeking Treatment for COVID-19 Infection.

|  | B | S.E. | aOR | 95% C.I. |  | Sig. |
| --- | --- | --- | --- | --- | --- | --- |
|  |  |  |  | Lower | Upper |  |
| ***COVID-19 Test and Symptom Status*** |  |  |  |  |  |  |
| COVID Test Negative and no Loss of Taste/Smell |  |  | Referent |  |  |  |
| COVID Test Positive and Loss of Taste/Smell | 1.341 | 0.184 | 3.823 | 2.626 | 5.567 | <.001 |
| COVID Test Positive and no Loss of Taste/Smell | 0.618 | 0.214 | 1.856 | 1.262 | 2.728 | 0.003 |
| COVID Test Negative and Loss of Taste/Smell | 1.196 | 0.214 | 3.306 | 2.086 | 5.241 | <.001 |
| ***Demographic Characteristics*** |  |  |  |  |  |  |
| Sex (Referent: Women) | -0.498 | 0.088 | 0.608 | 0.511 | 0.723 | <.001 |
| Age (per year) | -0.016 | 0.003 | 0.984 | 0.977 | 0.991 | <.001 |
| Marital Status (Referent: Married/Partner) | 0.212 | 0.087 | 1.236 | 1.042 | 1.046 | 0.015 |
| Race/Ethnicity |  |  |  |  |  |  |
| White |  |  | Referent |  |  |  |
| Black | 0.428 | 0.136 | 1.535 | 1.176 | 2.003 | 0.002 |
| Asian | 0.174 | 0.174 | 1.190 | 0.846 | 1.674 | 0.317 |
| Other Non-Hispanic | -0.184 | 0.193 | 0.832 | 0.569 | 1.215 | 0.340 |
| Hispanic | 0.290 | 0.096 | 1.337 | 1.107 | 1.615 | 0.003 |
| ***Anthropometric/Comorbidity Characteristics*** |  |  |  |  |  |  |
| Body Mass Index | -0.006 | 0.007 | 0.994 | 0.981 | 1.008 | 0.391 |
| Comorbidity | 0.146 | 0.020 | 1.158 | 1.112 | 1.205 | <.001 |
| *Number of COVID-19 Infections* |  |  |  |  |  |  |
| 0 |  |  | Referent |  |  |  |
| 1 | 0.215 | 0.264 | 1.240 | 0.651 | 2.360 | 0.447 |
| 2 | 0.436 | 0.259 | 1.547 | 0.846 | 2.830 | 0.133 |
| 3 | 0.559 | 0.319 | 1.748 | 0.874 | 3.499 | 0.105 |
| 4 | 0.810 | 0.379 | 1.085 | 0.507 | 2.321 | 0.831 |
| Exposed to COVID-19 | 0.386 | 0.107 | 1.471 | 1.192 | 1.816 | <0.001 |
| High Risk for COVID-19 | 0.161 | 0.113 | 1.174 | 0.940 | 1.467 | 0.156 |
| Symptoms Suggestive of COVID-19 | 0.670 | 0.159 | 1.954 | 1.431 | 2.667 | <.0001 |
| Current Insomnia (Referent: No insomnia) | 0.215 | 0.102 | 1.240 | 1.013 | 1.518 | 0.037 |
| Obstructive Sleep Apnea (Referent: No OSA) | 0.398 | 0.101 | 1.488 | 1.221 | 1.815 | <0.001 |
| ***Socioeconomic Characteristics*** |  |  |  |  |  |  |
| Income (Referent: <\$50,000) | 0.177 | 0.092 | 1.193 | 0.996 | 1.429 | 0.055 |
| Education (Referent: Some College) | 0.236 | 0.105 | 1.266 | 1.030 | 1.557 | 0.025 |
| Employment: (Referent: Full or Part-time, Student) | -0.005 | 0.131 | 0.995 | 0.763 | 1.297 | 0.967 |
| *Health Insurance* |  |  |  |  |  |  |
| Uninsured |  |  | Referent |  |  |  |
| Employer or Union | 0.218 | 0.245 | 1.243 | 0.769 | 2.011 | 0.375 |
| Self Purchase | 0.577 | 0.259 | 1.781 | 1.073 | 2.956 | 0.026 |
| Medicaid | 0.331 | 0.250 | 1.393 | 0.853 | 2.272 | 0.185 |
| Medicare | 0.835 | 0.253 | 2.305 | 1.405 | 3.782 | <0.001 |
| Other | 0.605 | 0.283 | 1.831 | 1.052 | 3.189 | 0.033 |
| *Political Preference (Referent: Moderate)* |  |  |  |  |  |  |
| Conservative | 0.266 | 0.127 | 1.304 | 1.017 | 1.672 | 0.036 |
| Liberal | 0.521 | 0.120 | 1.684 | 1.331 | 2.131 | <0.001 |
| Work Hours | 0.014 | 0.002 | 1.104 | 1.009 | 1.019 | <0.001 |

Table 2 contains aORs demonstrating that several demographic, socioeconomic, and comorbid factors are independently associated with seeking HAVT. Men, younger age, having a partner, and Black race or Hispanic ethnicity were associated with a higher probability of seeking HAVT. In addition, comorbidities including current insomnia and symptoms of OSA increased the likelihood of seeking HAVT. With respect to socioeconomic characteristics, higher educational attainment, self-purchase or Medicare health insurance and longer work hours were associated with greater probability of seeking HAVT. Both conservative and liberal political preference were related to seeking HAVT, but liberals were 1.33 times more likely than conservatives (aOR: 1.330, 95% CI: 1.150-1.538, not shown in Table 2).

In Table 3 are displayed the prevalences of factors potentially associated with receipt of HAVT. Similar to those seeking HAVT, persons who had lost their taste or smell or had a positive COVID-19 test, had a partner, were exposed to COVID-19, were at high risk for COVID-19, had symptoms suggestive of COVID-19, had higher income, were more educated, and who were either conservative or liberal were more likely to receive HAVT. However, age, BMI, race/ethnicity, comorbidity, number of COVID-19 infections, night shift work or work hours were not associated with a greater likelihood of receiving HAVT. In a logistic regression model (Table 4), only COVID-19 test and symptoms, having a marital partner, having employer or union supplied health insurance, and a liberal political affiliation were associated with receiving HAVT.

**Table 3:**
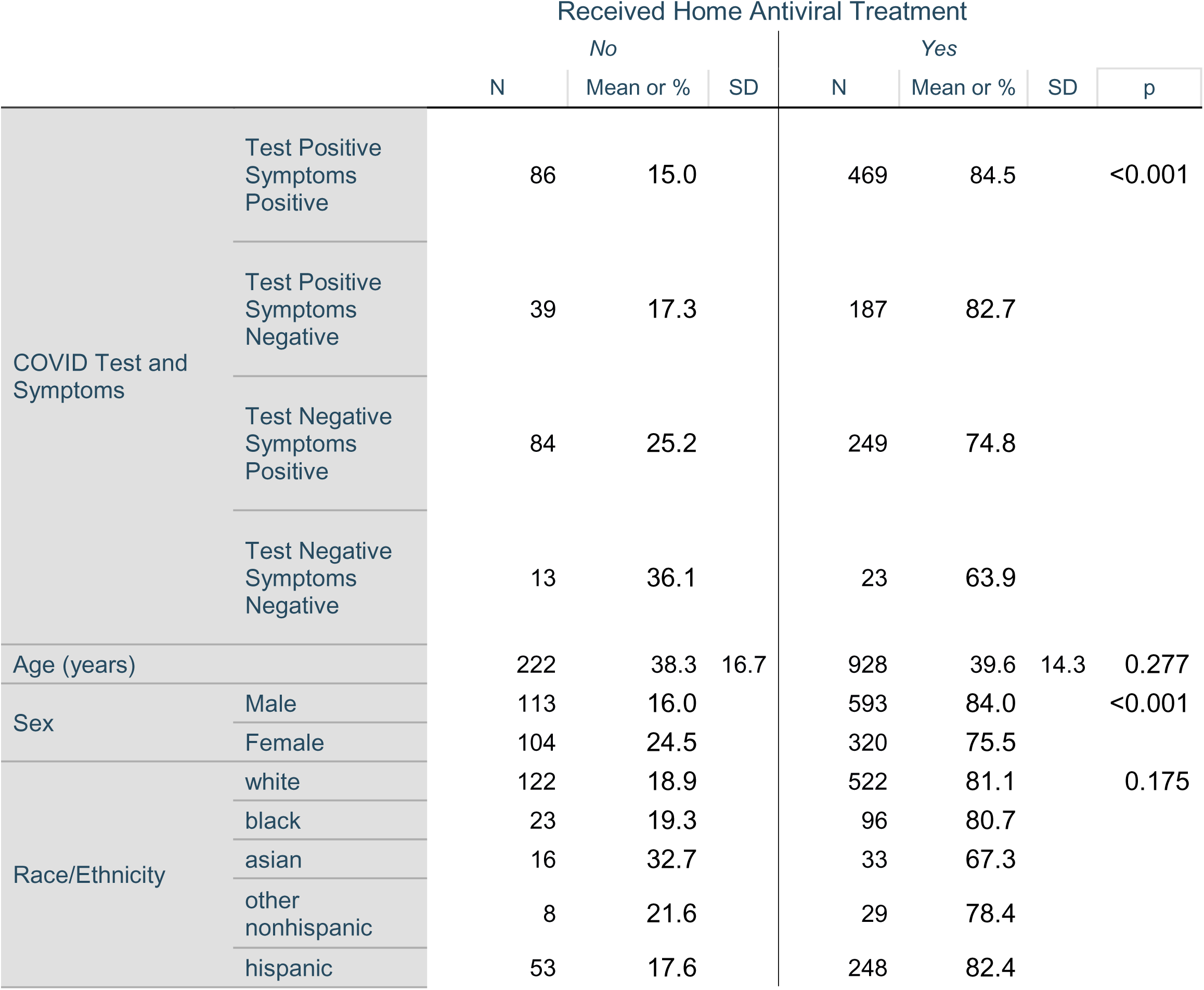
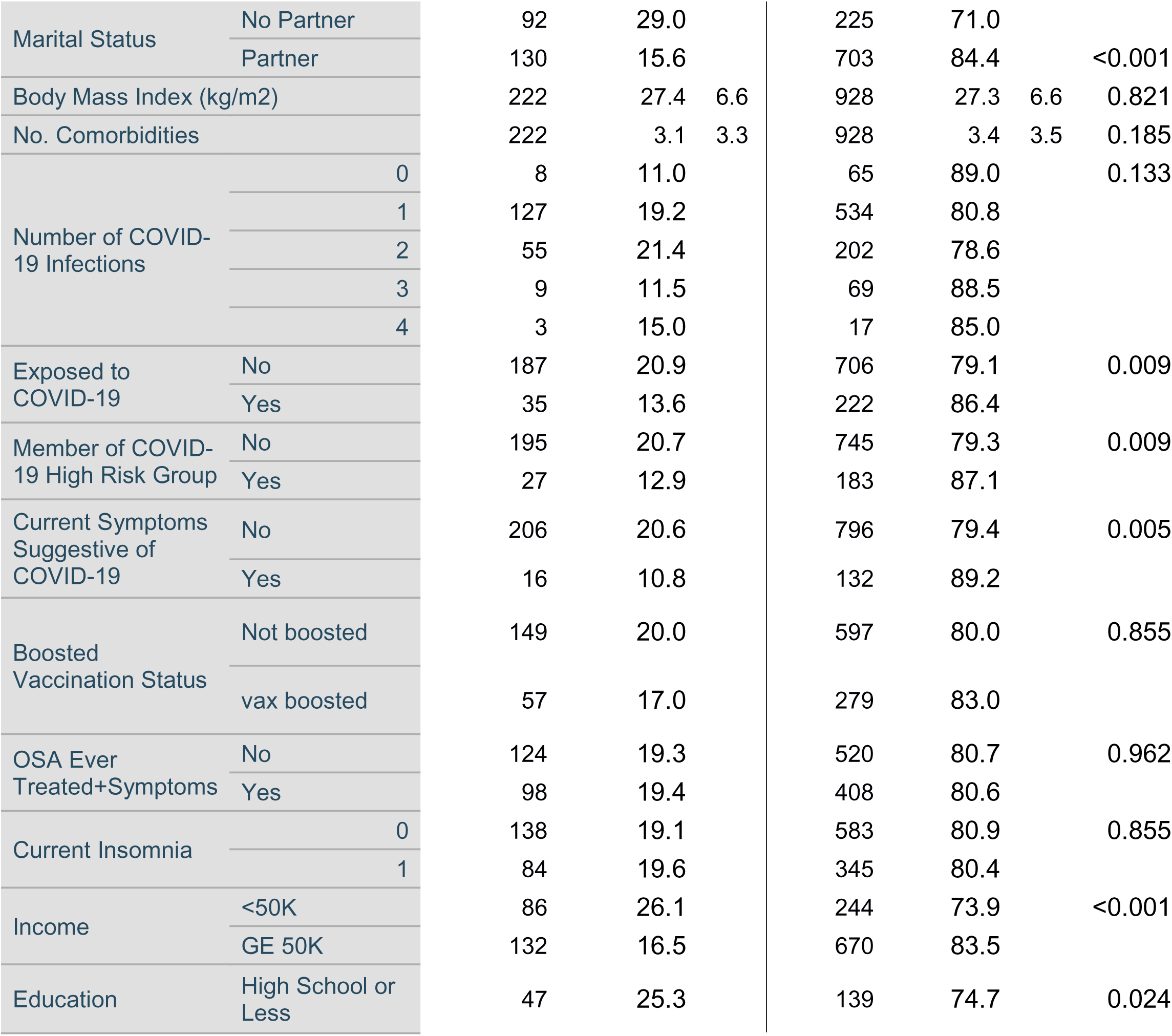
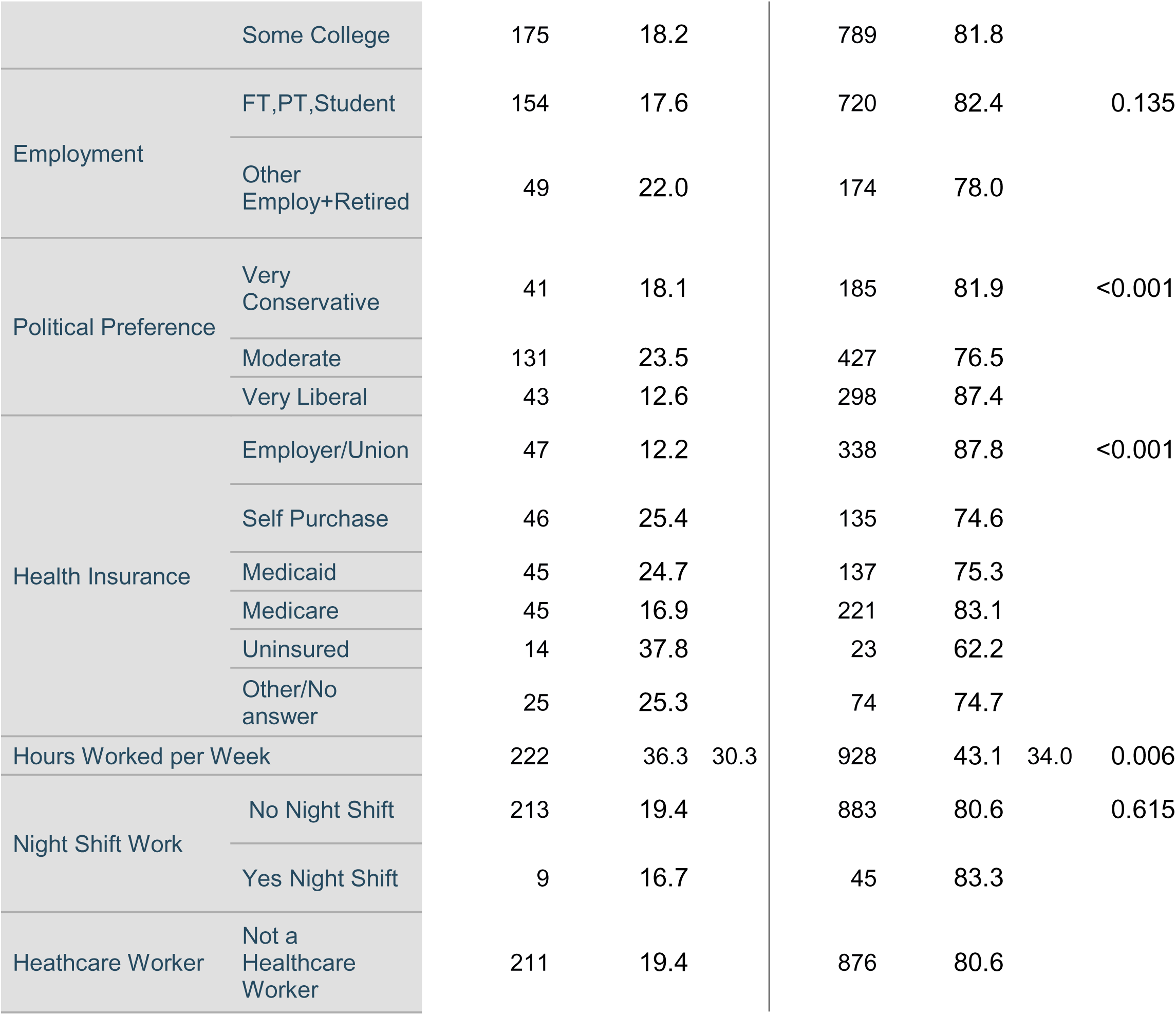
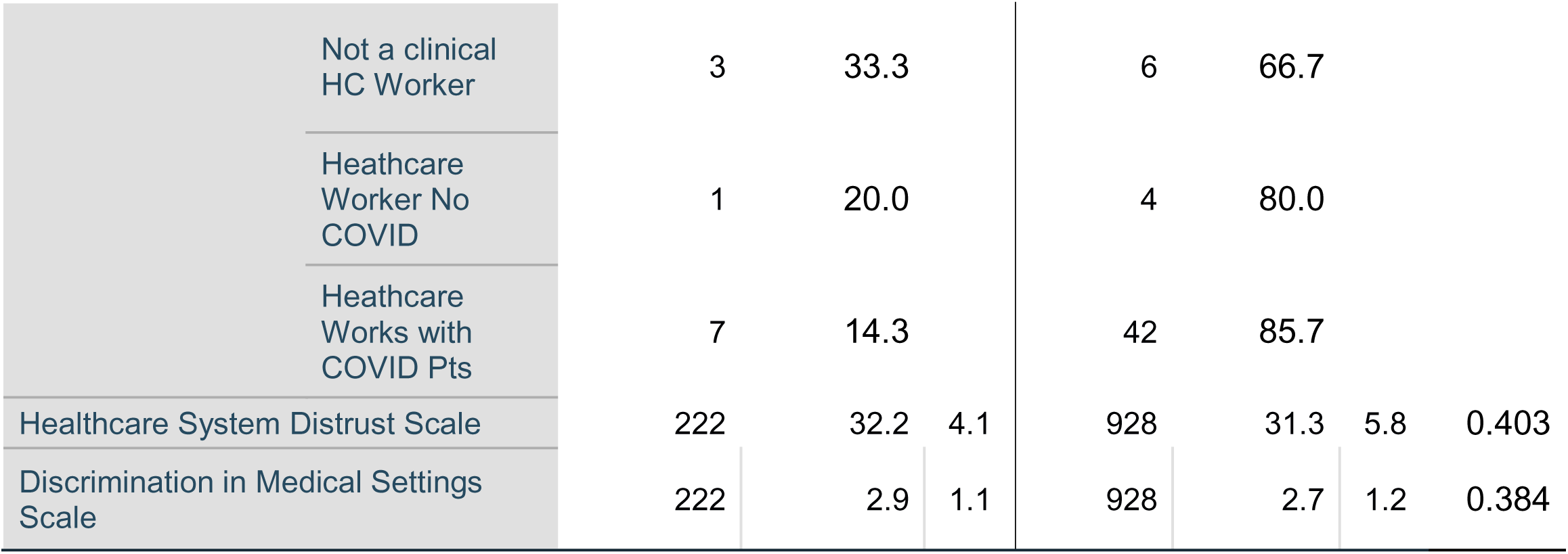
Factors Associated with Receiving Home Antiviral Treatment.

**Table 4:** Adjusted Odds Ratios of Factors Associated with Receiving Treatment for COVID-19 Infection.

|  | B | S.E. | aOR | 95% C.I. |  | Sig. |
| --- | --- | --- | --- | --- | --- | --- |
|  |  |  |  | Lower | Upper |  |
| **COVID-19 Test and Symptom Status** |  |  |  |  |  |  |
| COVID Test Negative and no Loss of Taste/Smell |  |  | Referent |  |  |  |
| COVID Test Positive and Loss of Taste/Smell | 0.992 | 0.465 | 2.698 | 1.022 | 7.120 |  |
| COVID Test Positive and no Loss of Taste/Smell | 1.023 | 0.512 | 2.782 | 0.943 | 8.208 |  |
| COVID Test Negative and Loss of Taste/Smell | 0.439 | 0.425 | 1.552 | 0.656 | 3.670 |  |
| **Demographic Characteristics** |  |  |  |  |  |  |
| Sex (Referent: Women) | -0.258 | 0.189 | 0.772 | 0.528 | 1.131 |  |
| Marital Status (Referent: Married/Partner) | 0.529 | 0.199 | 1.698 | 1.134 | 2.540 |  |
| Exposed to COVID-19 | 0.320 | 0.211 | 1.378 | 0.907 | 2.093 |  |
| High Risk for COVID-19 | 0.348 | 0.215 | 1.417 | 0.929 | 2.160 | 0.105 |
| Symptoms Suggestive of COVID-19 | 0.568 | 0.292 | 1.765 | 0.991 | 3.142 | 0.054 |
| **Socioeconomic Characteristics** |  |  |  |  |  |  |
| Income (Referent: <\$50,000) | 0.041 | 0.231 | 1.042 | 0.647 | 1.679 | 0.458 |
| Education (Referent: Some College) | 0.099 | 0.196 | 1.104 | 0.751 | 1.621 | 0.081 |
| **Health Insurance** |  |  |  |  |  |  |
| Uninsured |  |  | Referent |  |  |  |
| Employer or Union | 1.079 | 0.387 | 2.941 | 1.371 | 6.306 | 0.006 |
| Self Purchase | 0.226 | 0.415 | 1.254 | 0.549 | 2.864 | 0.588 |
| Medicaid | 0.371 | 0.388 | 1.449 | 0.673 | 3.119 | 0.341 |
| Medicare | 0.698 | 0.398 | 2.009 | 0.915 | 4.408 | 0.081 |
| Other | 0.388 | 0.404 | 1.474 | 0.667 | 3.259 | 0.337 |
| **Political Preference (Referent: Moderate)** |  |  |  |  |  |  |
| Conservative | 0.048 | 0.208 | 1.049 | 0.697 | 1.579 | 0.818 |
| Liberal | 0.622 | 0.198 | 1.863 | 1.262 | 2.751 | 0.002 |

In Table 5 are the influences of distrust in healthcare and medical discrimination on likelihood of seeking HAVT. In baseline models both medical discrimination and distrust in healthcare were associated with greater rates of seeking HAVT. However, in fully adjusted models, only medical discrimination retained statistical significance (aOR: 1.256, 95% CI: 1.108-2.424).

**Table 5:** Baseline and Adjusted Odds Ratios for the Association between Seeking Treatment for COVID-19 Infection and Medical Discrimination or Distrust in Healthcare.

|  | B | S.E. | OR/aOR | 95% C.I. for<br>OR/aOR |  | Sig. |
| --- | --- | --- | --- | --- | --- | --- |
| **Medical Discrimination** |  |  |  |  |  |  |
| Baseline | 0.634 | 0.045 | 1.877 | 1.719 | 2.050 | <0.001 |
| Fully Adjusted* | 0.228 | 0.064 | 1.256 | 1.108 | 2.424 | <0.001 |
| **Distrust Healthcare** |  |  |  |  |  |  |
| Baseline | 0.061 | 0.008 | 1.062 | 1.047 | 1.078 | <0.001 |
| Fully Adjusted* | 0.003 | 0.010 | 1.003 | 0.984 | 1.022 | 0.771 |

\*Adjusted for COVID-19 Test and Symptom Status, sex, age, marital status, race/ethnicity, body mass index, comorbidity, number of COVID-19 infections, exposure to COVID-19, high risk for COVID-19, symptoms suggestive of COVID-19, current insomnia, obstructive sleep apnea, income, education, employment, health insurance, political preference, work hours

Table 6 lists the prevalence of various barriers to obtaining treatment. The most common was feeling uncomfortable obtaining the medication while being sick (n=47, 3.5%) while the least common was having a contraindication to the medication (n=2, 0.2%). Overall, 14.7% of those who sought HAVT documented at least one barrier to obtaining treatment.

**Table 6:** Barriers to Home Antiviral Treatment for COVID-19.

Number Seeking Treatment: 1331
|  | *N* | % |
| --- | --- | --- |
| No Medication Nearby | 33 | 2.3 |
| No Transportation Available | 43 | 3.2 |
| Uncomfortable Obtaining While Sick | 47 | 3.5 |
| Didn't Receive the Prescription Quickly Enough | 33 | 2.5 |
| Didn't Want to be Charged for the Medication | 29 | 2.2 |
| Could not Locate Pharmacies with the Medication | 26 | 2 |
| Did not Think it would be Effective | 43 | 3.2 |
| Symptoms Not Sufficiently Severe | 43 | 3.2 |
| Physician Would Not Prescribe | 5 | 0.4 |
| Contradicated Due to Other Medical Issues | 2 | 0.2 |
| At Least 1 Barrier to Obtaining Treatment | 195 | 14.7 |

## Discussion

In this analysis performed in a large, diverse general U.S. adult population cohort, 26.3% of those who believed they had a COVID-19 infection sought HAVT and only 69.7% of those who sought it actually received the medication. Furthermore, the factor most strongly associated with seeking HAVT was experiencing a loss of taste or smell. Although several other demographic and socioeconomic factors, and medical comorbidities also were associated with seeking HAVT, it is notable that the likelihood in persons of Black race and Hispanic ethnicity was greater than Whites. In addition, experiencing discrimination while receiving healthcare was associated with a higher rate of seeking HAVT. In contrast, racial, ethnic and income disparities were not observed among those receiving HAVT.

Our finding that a significant minority of persons potentially eligible for COVID-19 HAVT seek the medication and even fewer receive it is consistent with several studies using data from EMRs and other databases [9,11,13]. An EMR analysis conducted in a large healthcare organization found that the proportion of treatment eligible receiving nirmatrelvir–ritonavir increased from 1.1% at the beginning of 2022 to 24.0% at the end of the year [9]. This is consistent with the 21.3% of persons who received HAVT in our general population cohort. Furthermore, a retrospective analysis of EMR data from the National COVID Cohort Collaborative found that nirmatrelvir–ritonavir use increased from 1.2% at the beginning of 2022 to 35.1% at the end of 2023 [26]. To our knowledge, although indications for use of HAVT have expanded, there is no current data regarding the percentage of COVID-19 infected persons who seek HAVT. During the pandemic, the federal government [3] as well as drug manufacturers and various non-governmental organizations [27,28] attempted to increase public awareness of the availability of HAVT. Nevertheless, public awareness remained low which likely contributed to lack of utilization [29]; it still remains low today [8]. Inasmuch as HAVT was free during the time interval of our survey, the lack of public awareness is illustrated further by our finding that those who were uninsured had the lowest rates of seeking HAVT. Additionally, we identified a number of barriers that hindered access. Although lack of HAVT availability should no longer be a barrier since medication is now in sufficient supply, there are fewer pharmacies located in socioeconomically disadvantaged areas [30]. This will contribute to difficulty accessing HAVT especially if the infected individual is ill. Furthermore, there is a large amount of misinformation regarding HAVT with posts on social media questioning its effectiveness and advocating for disproven and dangerous alternatives [31,32]. Currently for most persons with COVID-19 infection, symptoms are relatively mild, but this may contribute to lack of interest in HAVT [13]. However, progression to severe illness still occurs especially among those at high risk such as the elderly [33], and importantly is mitigated with the use of HAVT [34]. Collectively, evidence thus suggests that underutilization of HAVT is multifactorial and related to lack of awareness, structural inequities in the healthcare system and misinformation regarding effectiveness.

The most important factor in this study associated with seeking HAVT was the loss of taste or smell. The aOR for a positive COVID-19 test and loss of taste or smell was over twice that of only having a positive COVID-19 test (3.823 vs. 1.856). As recently reviewed, loss of smell or taste has been noted in some studies to be present in more than 80% of persons with COVID-19 [35] although the incidence of these symptoms may be much lower with the Omnicron variants of the virus [36]. Its presence has become a marker of COVID-19 in contradistinction to its rarity in influenza and other respiratory illnesses [37]. As a result of public media reporting, public awareness of its association with COVID-19 also is high [38]. Other symptoms of COVID-19 such as fever or sore throat are non-specific and are not as likely to motivate individuals to seek evaluation and treatment for COVID-19. This is exemplified in our study by a lower aOR in response to a question associated with “experiencing symptoms concerning for COVID-19” that did not include loss of taste or smell as an example. Therefore, a public awareness campaign highlighting the linkage between loss of taste or smell and COVID-19 may be a means to increase usage of HAVT.

Our findings from a general US population that men, younger age, Black race and Hispanic ethnicity are associated with a higher likelihood of seeking HAVT are inconsistent with previous reports that older persons, Whites and Asians have higher rates of HAVT being dispensed [9–12,15]. This suggests that assessment of dispensing data may be misleading with respect to the motivation of different population groups in seeking HAVT. The disparity between seeking and receiving (dispensing) HAVT also is highlighted in our current analysis in which we did not observe sex, racial or ethnic differences in receipt of HAVT. In contrast, the presence of loss of taste or smell, or a positive COVID-19 test were strongly associated with receipt of HAVT.

In contrast to studies showing lower rates of dispensing HAVT in areas with high social vulnerability [5,14,39], we found that seeking HAVT was associated with higher metrics of socioeconomic status as reflected by educational attainment, income and employment. These differences were no longer apparent among those who received HAVT in our study, yet they nevertheless reinforce the concept that factors associated with seeking HAVT are different than those associated with receiving it. Therefore, developing measures to increase utilization of HAVT will need to consider both outcomes.

We observed that in comparison to those with moderate political preferences, both conservatives and liberals were more likely to seek HAVT. During the pandemic, substantial misinformation regarding treatment of COVID-19 was promulgated by right-wing social media as well as by the conservative leaders of the federal government [32]. Thus, it is surprising that conservatives were only slightly less motivated than liberals to seek HAVT. However, as recently reviewed, social psychological research indicates that conservatism is associated with threat such as disease and that socio-ecological data suggest that conservatism also is related to higher levels of communicable disease [40]. Thus, it is possible that these underlying beliefs outweighed the effects of partisan messaging as they relate to seeking HAVT. However, this was not the situation when receiving HAVT, for reasons that are unclear.

Both distrust in the healthcare system and experiencing discrimination when receiving medical care are associated with a decrease in seeking medical care [21,41]. However, our data paradoxically found that medical discrimination but not healthcare distrust was related to a greater likelihood of seeking HAVT. The explanation is unclear, but reverse causality is possible; the act of seeking HAVT engendered more medical discrimination. This finding requires additional investigation.

There are several limitations of our study and analyses. First, all of the data regarding COVID-19 infection, HAVT and the various covariates are self-reported. Thus, they are subject to recall bias resulting in misclassification of outcomes and exposures. Second, the number of participants in our cohort is relatively small in comparison to studies assessing dispensing of HAVT. Third, data collection for this study occurred 8-10 months after availability of HAVT and is likely not representative of the initial introduction of HAVT or current utilization. Nevertheless, it should be emphasized that to our knowledge, this is the only study in a diverse general population that has assessed factors associated with seeking and receiving HAVT.

In conclusion, our study provides robust evidence that HAVTs for COVID-19 are an underutilized measure to reduce the risk of severe COVID-19 among U.S. adults. Seeking and receiving HAVT is highly linked to the presence of loss of taste or smell, more so than other symptoms. Several other demographic and socioeconomic factors also are associated with seeking HAVT and these appear to be different than those associated with receiving HAVT. Our data suggest that factors related both seeking and receiving HAVT need to be considered in designing future public awareness campaigns to increase utilization particularly in high risk populations.

**Figure:**
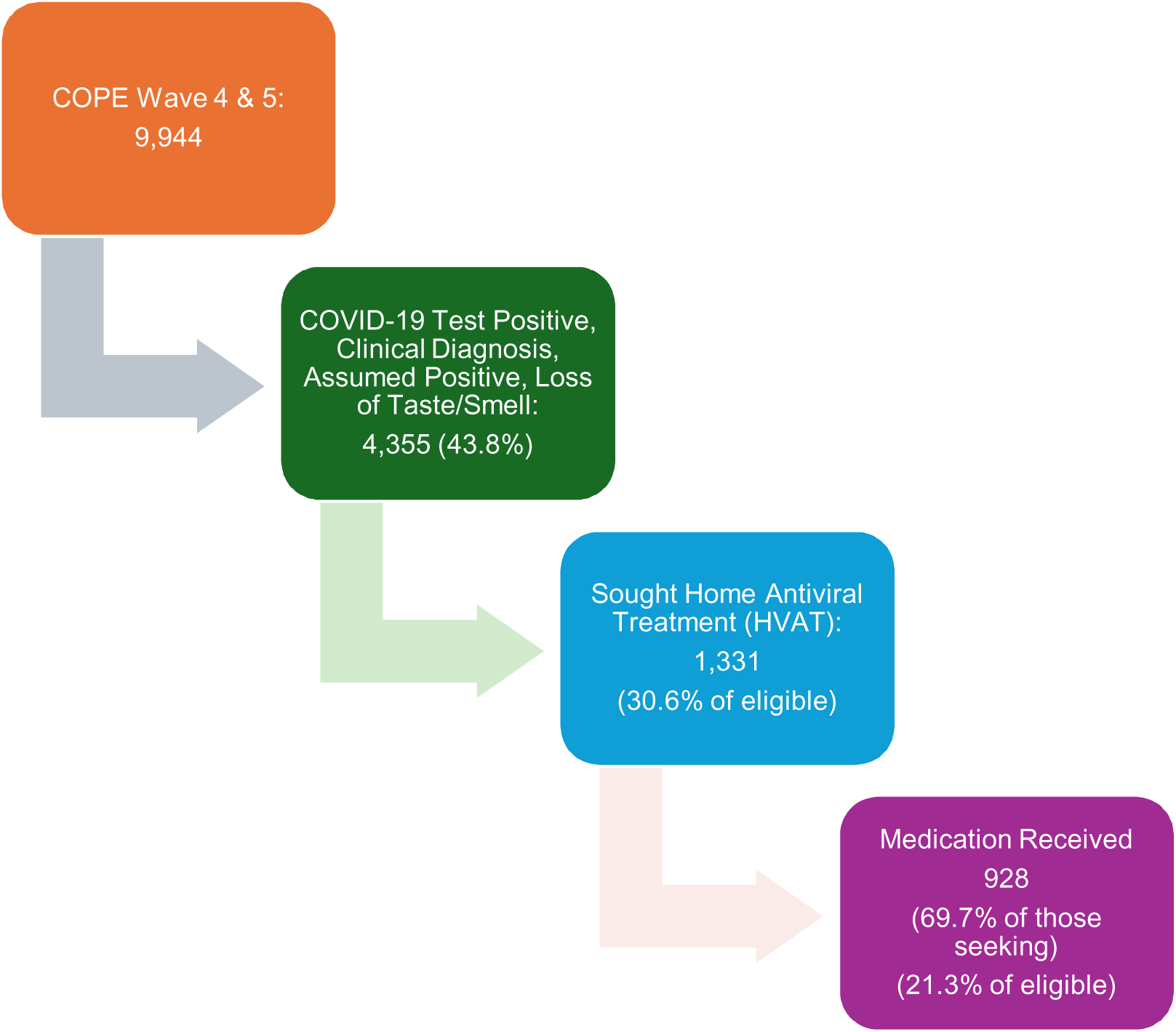
Flow chart of included participants

## Declarations

### Ethics Approval

All procedures were in accordance with the ethical standards of Monash University Human Research Ethics Committee (Study #24036) and with the 1964 Helsinki declaration and its later amendments or comparable ethical standards. Informed consent was obtained electronically from all individual participants included in the study.

### Author Approval

All authors have seen and approved the manuscript.

### Acknowledgments of Author Contributions

Concept and Design: SFQ

Data collection: MDW, MÉC, MEH

Data analysis and interpretation: SFQ

Drafting of the manuscript: SFQ

Critical feedback and revision of manuscript: SFQ, MDW, MÉC, LAB, MEH, MLJ, CFM, AR, RR, PV, SMWR, CAC

### Data Availability

The data that support the findings of this study are available from the corresponding author upon reasonable request subject to any institutional review board constraints.

