## Supplementary material for "Factors Associated with Seeking and Receiving Home Antiviral Therapy During the COVID-19 Pandemic": Methods Supplement

Supplement 1: Additional Demographic Survey Questions

What is the **primary** source of your health coverage?

- A plan purchased through an employer or union (1)
- A plan that you or another family member buys on your own (2)
- Medicare (4)
- Medicaid or other state program (5)
- Some other source (6)
- No health coverage (7)
- Prefer not to answer (8)

What is your political affiliation? (Select one)
 *Note: In modern US politics, normally Democratic Party ideals are considered liberal and Republican Party ideals are considered conservative.*

|  | Very Liberal (1) | Slightly Liberal (2) | Neither Liberal nor Conservative (3) | Slightly Conservative (4) | Very Conservative (5) | Prefer not to answer (6) |
| --- | --- | --- | --- | --- | --- | --- |
| I consider myself... (1) |  |  |  |  |  |  |

In the past **week**, how many total **hours** did you spend doing *paid* work...

|  | Hours per week |
| --- | --- |

|  | 0 | 20 | 40 | 60 | 80 | 100 | 120 | 140 |
| --- | --- | --- | --- | --- | --- | --- | --- | --- |

| 1 () | 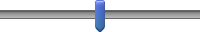 |
| --- | --- |

**Currently**, about how often do you work a night shift (i.e., the majority of your work hours occur between 10pm and 8am)? *(Select one)*

- Nearly every day (1)
- 3-4 times per week (2)
- 1-2 times per week (3)
- 1-2 times per month (5)
- Never or nearly never (6)

In what area or type of business is this job/occupation?

If healthcare is selected:

In your role as a healthcare provider, do you have direct patient contact?

- Yes, including the treatment of diagnosed COVID-19 patients (1)
- Yes, but not including the treatment of diagnosed COVID-19 patients (2)
- No (3)
