## Supplementary material for "Factors Associated with Seeking and Receiving Home Antiviral Therapy During the COVID-19 Pandemic": Table S1

| **Table S1: Unadjusted Odds Ratios of Factors Associated with Seeking Treatment for COVID-19 Infection** | | | | | |  |
| --- | --- | --- | --- | --- | --- | --- |
|  | **B** | **S.E.** | **Exp(B)** | **95% C.I.for EXP(B)** | | **Sig.** |
|  |  |  |  | **Lower** | **Upper** |  |
| ***COVID-19 Test and Symptom Status*** |  |  |  |  |  |  |
| COVID Test Negative and no Loss of Taste/Smell |  |  | Referent |  |  |  |
| COVID Test Positive and Loss of Taste/Smell | 1.839 | 0.165 | 6.29 | 4.549 | 8.697 | <.001 |
| COVID Test Positive and no Loss of Taste/Smell | 0.54 | 0.17 | 1.717 | 1.231 | 2.394 | 0.001 |
| COVID Test Negative and Loss of Taste/Smell | 1.732 | 0.17 | 5.654 | 4.053 | 7.886 | <.001 |
| ***Demographic Characteristics*** |  |  |  |  |  |  |
| Sex (Referent: Men) | -0.804 | 0.068 | 0.447 | 0.392 | 0.511 | <.001 |
| Age (per year) | -0.02 | 0.002 | 0.98 | 0.976 | 0.984 | <.001 |
| Marital Status (Referent: No Partner) | 0.494 | 0.071 | 1.639 | 1.428 | 1.883 | <.001 |
| Race/Ethnicity |  |  |  |  |  |  |
| White |  |  | Referent |  |  |  |
| Black | 0.283 | 0.114 | 1.327 | 1.06 | 1.66 | 0.013 |
| Asian | -0.033 | 0.159 | 0.968 | 0.709 | 1.321 | 0.836 |
| Other Non-Hispanic | -0.568 | 0.175 | 0.567 | 0.402 | 0.798 | 0.001 |
| Hispanic | 0.322 | 0.079 | 1.38 | 1.182 | 1.613 | <.001 |
| ***Anthropometric/Comorbidity Characteristics*** |  |  |  |  |  |  |
| Body Mass Index | -0.014 | 0.006 | 0.986 | 0.975 | 0.997 | 0.014 |
| Comorbidity | 0.288 | 0.014 | 1.334 | 1.299 | 1.371 | <.001 |
| *Number of COVID-19 Infections* |  |  |  |  |  |  |
| *0* |  |  | Referent |  |  |  |
| 1 | -0.558 | 0.137 | 0.572 | 0.438 | 0.748 | <.001 |
| 2 | 0.008 | 0.15 | 1.008 | 0.752 | 1.352 | 0.957 |
| 3 | 0.383 | 0.199 | 1.466 | 0.993 | 2.164 | 0.054 |
| 4 | 0.101 | 0.318 | 1.106 | 0.593 | 2.065 | 0.751 |
| Exposed to COVID-19 | 0.804 | 0.087 | 2.235 | 1.884 | 2.653 | <.001 |
| High Risk for COVID-19 | 0.254 | 0.088 | 1.289 | 1.084 | 1.533 | 0.004 |
| Symptoms Suggestive of COVID-19 | 1.554 | 0.136 | 4.731 | 3.626 | 6.173 | <.001 |
| Current Insomnia (Referent: No insomnia) | 0.94 | 0.074 | 2.56 | 2.214 | 2.96 | <.001 |
| Obstructive Sleep Apnea (Referent: No OSA) | 1.41 | 0.075 | 4.095 | 3.535 | 4.743 | <.001 |
| ***Socioeconomic Characteristics*** |  |  |  |  |  |  |
| Income (Referent: <$50,000) | 0.648 | 0.07 | 1.912 | 1.667 | 2.193 | <.001 |
| Education (Referent: Some College) | 0.516 | 0.083 | 1.675 | 1.424 | 1.97 | <.001 |
| Employment: (Referent: Full or Part-time, Student) | -0.929 | 0.078 | 0.395 | 0.339 | 0.46 | <.001 |
| *Health Insurance* |  |  |  |  |  |  |
| Uninsured |  |  | Referent |  |  |  |
| Employer or Union | 0.844 | 0.173 | 2.326 | 1.658 | 3.262 | <.001 |
| Self Purchase | 1.143 | 0.186 | 3.137 | 2.179 | 4.518 | <.001 |
| Medicaid | 0.649 | 0.181 | 1.913 | 1.342 | 2.726 | <.001 |
| Medicare | 0.896 | 0.177 | 2.451 | 1.733 | 3.467 | <.001 |
| Other | 0.912 | 0.199 | 2.489 | 1.684 | 3.679 | <.001 |
| *Political Preference (Referent: Moderate)* |  |  |  |  |  |  |
| Conservative | 0.471 | 0.09 | 1.602 | 1.343 | 1.91 | <.001 |
| Liberal | 1.024 | 0.085 | 2.785 | 2.358 | 3.288 | <.001 |
| Work Hours | 0.024 | 0.001 | 1.024 | 1.022 | 1.027 | <.001 |
| Night Shifts | 0.346 | 0.161 | 1.413 | 1.031 | 1.938 | 0.032 |
| Distrust Healthcare | 0.061 | 0.008 | 1.063 | 1.047 | 1.079 | <.001 |
|  | 0.626 | 0.045 | 1.869 | 1.71 | 2.043 | <.001 |
